# The association between cardiovascular health and peripheral and central auditory functions in adults: a protocol for a systematic review

**DOI:** 10.1101/2022.08.13.22278662

**Authors:** Rosie C. Daly, Emma O’Donnell, Laura Barrett, Christian Füllgrabe

**Affiliations:** School of Sport, Exercise and Health Sciences, Loughborough University, Loughborough, United Kingdom; Ear Institute, University College London, 332 Gray’s Inn Road, London WC1X 8EE, United Kingdom

**Keywords:** auditory functions, cardiovascular health, speech identification, hearing loss, rehabilitation strategy

## Abstract

**Introduction:** The ability to process sounds decreases with advancing age and the already high prevalence of people with hearing loss (HL) is estimated to increase further over time. Hearing loss reduces speech identification which is important for day-to-day communication. In addition, it can lead to social isolation, depression, and lower quality of life. Current hearing rehabilitation strategies (eg, hearing aids) provide some benefits, but are not always accepted by hearing-impaired listeners and are less successful in real-life listening situations. Consequently, alternative rehabilitation strategies, such as the manipulation of cardiovascular (CV) health for the prevention and rehabilitation of HL, should be explored. Some research suggests that CV health and auditory functions are related, but the existence of such a link has not been systematically evaluated. This manuscript outlines the protocol for a systematic review of published research on the association between CV health and peripheral and central auditory functions across the adult lifespan and for all levels of hearing abilities.

**Method and analysis:** The Preferred Reporting Items for Systematic reviews and Meta-Analyses Protocols (PRISMA-P) checklist will be followed. Studies included for analysis will be original peer-reviewed articles, measuring cardiovascular health and hearing abilities to explore their relationship. Participants will be aged ≥18 years and will have various levels of hearing sensitivity and of CV health. Databases will be searched, using key words, to obtain evidence that meets the defined set of inclusion criteria. Data will be extracted and examined by two reviewers. Quality checks will occur, and, if appropriate, a meta-analysis will be performed. Data analysis will be completed and reported in a full systematic review, following the PRISMA guidelines.

**Ethics and dissemination:** No ethical approval is needed for the systematic review as only published data will be analysed. Findings will be disseminated at conferences and in peer-reviewed journals.

**Systematic review registration:** PROSPERO CRD42022353002

**STRENGTHS AND LIMITATIONS OF THIS STUDY:**

- The protocol follows the guidelines set out in the Preferred Reporting Items for Systematic Review and Meta-Analysis Protocols (PRISMA-P).
- The systematic review will consider both direct and indirect measures of CV health.
- The relationship of CV health with peripheral and with central auditory functioning will be examined.
- Results will indicate whether the manipulation of CV health could be used as an alternative rehabilitation strategy for HL.
- The systematic review will only include studies in the English language.

## INTRODUCTION

Hearing loss (HL) is generally defined as a reduction in hearing sensitivity, with pure-tone audiometric thresholds equal to or worse than ∼20 dB Hearing Level across a frequency range important for speech identification. In 2019, it was estimated that 1.6 billion people worldwide had some degree of HL,[1] and the prevalence of HL is predicted to rise to just under 2.5 billion by 2050.[2] Presbycusis (ie, age-related sensorineural HL) is the most common form of HL. It is already noticeable in middle age[3] and its severity increases with age.[4]

Hearing loss can be very debilitating for the affected individual. It results in reduced speech identification, especially in the presence of background noise that often occurs in everyday, social listening settings (such as a busy restaurant).[5] People with HL also experience more loneliness and social isolation, and report a lower quality of life than normal-hearing (NH) individuals.[6-8] Furthermore, HL is also associated with poor mental health, such as low mood and depression,[9] with individuals with HL being twice as likely to use anti-depressants and anti-anxiety medications.[10] Additionally, HL has been suggested to lead to lower cognitive performance[11] (for a critical discussion, see Füllgrabe, 2020)[12] and to the acceleration of cognitive decline that is normally associated with aging.[13] Recently, it was reported that HL is the largest modifiable mid-life risk factor for dementia.[14]

In addition to the psychosocial impact on the hearing-impaired (HI) individual, HL also imposes a large financial burden on society. In 2019, Europe’s healthcare economic expenditure for individuals with HL was approximately £24 million.[15] It has been noted that HI individuals use primary and secondary healthcare services more often than NH individuals.[16]

In some countries, such as Australia and the United States of America, the cost of hearing healthcare can also have a direct financial impact on the HI individual as hearing healthcare services are not necessarily covered by health insurance.[17] Having to pay for these services can constitute a barrier to accessing hearing healthcare, especially for those from low socioeconomic backgrounds.[18]

Hearing loss also has an indirect economic impact on society in terms of lower productivity in the workplace. Compared to NH individuals, individuals with HL perceive their work as more demanding.[19, 20] Additionally, HI employees report higher levels of fatigue and mental distress, which are also given more frequently as reasons for sick leave and absence from work than by NH employees.[21]

Different rehabilitation strategies have been developed to improve hearing abilities of HI individuals, and to lower the prevalence of negative side effects of HL. Currently, the standard treatment for mild-to-moderately-severe HL is the provision of hearing aids (HAs).[22] These devices provide frequency-specific amplification with the aim of making inaudible sounds at least partially audible again. However, the uptake of HAs remains low;[23] for example, in the United Kingdom, only up to 40% of HA owners wear their devices.[24] Possible reasons for this include: (i) the anticipated or experienced age-related stigma associated with the use of HAs;[25] (ii) the decline in dexterity with age, affecting the users’ ability to manipulate the HAs;[26] and (iii) the discomfort caused by wearing HAs (eg, skin irritations).[27] Probably most importantly, older HA users frequently complain about persisting difficulties with speech-in-noise (SiN) identification, despite receiving appropriate amplification from their HAs.[28, 29]

Auditory training programmes have been developed (such as Listening and Communication Enhancement Auditory Training; LACE) to help speech understanding in people with HL. There is evidence that such programmes do indeed improve performance on the trained speech tasks.[30] However, only minor improvements in speech identification have been demonstrated for untrained speech materials, meaning that the benefits derived from the auditory training do not generalise much to other listening situations.[31]

Another rehabilitation strategy that, this time, targets directly the causes of HL is gene therapy, which is based on suppressing and modifying the affected gene(s) that are involved in HL.[32] While this is a promising approach that has been successfully tested in animals, gene therapy is still in the early stages of its development and has yet to be applied to humans. It is also predicted to be relatively costly.[33]

Taken together, the before-mentioned rehabilitation strategies have been shown to provide benefits to HI individuals, but they also have clear limitations and drawbacks. Therefore, it is important to explore new clinically significant and cost-effective rehabilitation strategies for people with HL.

One possible alternative approach is via the cardiovascular (CV) system. Indeed, there is seemingly converging evidence that auditory functions and CV health are associated.[34-39] Furthermore, a causal relationship between CV health and auditory functions has been demonstrated by improvements in CV health following a physical-exercise program, resulting in better peripheral hearing functions.[40] However, not all studies investigating CV health and auditory functions found evidence in favour of the existence of a link between the two.[41-45]

The observed discrepancy between study findings could be explained by the different measures used to assess CV health. Some studies used ‘direct’ measures of CV health (eg, maximum amount of oxygen consumption during exercise, arterial stiffness)[34, 38] while others used ‘indirect’ measures, capturing the risk factors of CV disease (eg, self-reported history of smoking or hypertension).[43, 44] In addition, the ages and age range of the study participants were not consistent across studies (young vs older adults).[40, 42] Similarly, the hearing status of the participants and the range of included HL often varied (NH vs HL).[35, 43] However, the existence and strength of the association between CV health and auditory functions may differ across adulthood and with changes in hearing abilities.[46]

It is noteworthy that all research on the association between CV health and hearing abilities (with the exception of Goderie et al., 2021)[47] seemed to have focused on peripheral auditory functions as indexed, for example, by audiometric thresholds or otoacoustic emissions.[48] This is somewhat surprising as audiometric sensitivity is not a strong predictor of the ability to understand speech in interfering backgrounds,[49, 50] which is however the main complaint of HI listeners.[51] The association between CV health and supra-threshold (spectral, temporal and spatial) auditory processing abilities also has not been explored. However, these processing abilities contribute to successful speech perception,[52, 53] and are at least partially independent of audiometric loss.[54-56] This raises the question of whether, so far, research has focused on the most relevant outcome measures when exploring the association between CV health and auditory functions.

When investigating the impact of CV health on auditory functioning, it is of interest to consider the mediating effects of cognition in this relationship, as cognitive abilities are also associated with CV health[57, 58] and have been shown to play a role in a variety of supra-threshold auditory processing abilities (especially SiN identification).[52, 59]

Several efforts have been made to overview the literature on the impact of CV health on hearing. Some authors focused on the role of CV health in the vulnerability to noise and in noise-induced HL,[60] while others looked more broadly at the association between CV health and HL. As regards the latter category, for example, Besser et al. (2018)[61] reviewed the literature to recommend adaptations of hearing healthcare by considering multimorbidity, and Yildirim (2012)[62] and Oron et al. (2014)[63] examined the evidence that CV health is a potential cause of HL. However, none of these publications followed established guidelines for conducting systematic reviews (eg, Preferred Reporting Items for Systematic reviews and Meta-Analyses, PRISMA 2020).[64] Hence, it is not certain that all relevant studies were included in these reviews of the literature.

Some systematic reviews can be found in the literature. However, the studies included in these systematic reviews often used narrow age ranges and specific types of HLs (in terms of their severity and etiology). Hence, it is unknown whether the conclusions of these systematic reviews are applicable across the adult lifespan and to all hearing abilities. For example, some systematic reviews focused on sudden sensorineural HL,[65, 66] leaving it unclear whether CV health also plays a role in the more prevalent age-related HL. Other systematic reviews focused on data obtained from children and adolescents,[67] thus excluding the age group for which HL is most prevalent (ie, older adults). In addition, many systematic reviews focused on studies using a single (and generally indirect) indicator of CV health.[68, 69]

In summary, to date, there has been no attempt to comprehensively characterise the relationship between CV and auditory health while differentiating the adult study populations in terms of age and hearing ability on the one hand, and CV-health and auditory measures on the other. The proposed systematic review will evaluate the evidence for the impact of CV health on peripheral and central auditory functions from studies covering all levels of hearing abilities in adults drawn from across the entire adult lifespan. Both indirect and direct CV-health measures will be considered. The findings of this systematic review will help to determine if CV health plays a role in auditory health, and, thus, can be targeted for the prevention and rehabilitation of HL.

## METHODS AND ANALYSIS

The protocol for the systematic review will follow the guidelines of the Preferred Reporting Items for Systematic Review and Meta-Analysis-Protocol (PRISMA-P) 2015 checklist.[70] The systematic review will follow the guidelines of the PRISMA 2020 checklist.[64] The systematic review will be led by author RD.

### Eligibility criteria

The inclusion and exclusion criteria for the systematic review were created based on an approach of population, factors and outcomes (PFO).[71] Each is described below, in addition to the types of studies that will be searched.

#### Types of study

Only original studies that have been published in peer-reviewed journals will be included. Editorials, opinion pieces, reviews, individual case studies, conference and poster abstracts, and book chapters will be excluded unless they include all appropriate original data and have been published and peer reviewed. Studies whose full text is not available in English will be excluded.

#### Population

Studies with adult participants aged 18 years or above (ie, with no maximum age) will be included. Studies also using younger (<18 years of age) participants will only be included if the results for the adult participants are reported separately. Studies using participants with a confirmed conductive HL will be excluded. Studies will not be excluded based on their participants’ geographical location, sex, or race/ethnicity. Studies using non-human participants will be excluded.

#### Factors

Studies included must report on at least one aspect of CV health that may influence hearing, based on direct measures of CV health (eg, the cardiorespiratory fitness, arterial function and structure) or indirect measures of CV health [eg, CV risk factors such as body mass index (BMI), self-report of smoking status, diabetes and hypertension].

#### Outcomes

Studies included must report on at least one aspect of auditory functioning, based on peripheral auditory measures (eg, audiometric thresholds, OAEs) or central auditory measures (eg, speech-identification tasks). Studies in which participants self-reported their hearing status or speech-identification abilities will also be included.

To be included in this systematic review, studies must have recorded both cardiovascular health and hearing abilities and explored their relationship. Studies that investigated only the impact of auditory functions on CV health will not be included.

### Information sources

The literature search will be conducted on multiple databases across the disciplines of psychology, medicine and physiology to identify appropriate studies. The following databases will be searched: PubMed, Embase, PsychArticles, PsychINFO, Scopus, Web of Science and Wiley Online Library. An initial search through all databases will be carried out on the same day, using an established list of key words and, where appropriate, Medical Subject Headings (MeSH) terms. There will be no restrictions on publication date, language or type. In addition, the reference lists of studies published in the four months preceding the literature search and the work of key authors (defined as authors whose names appear as a first author more than five times in the search results) will be manually searched for any published articles not found during the initial search. The number of studies retrieved as a result of the initial and additional searches will be presented in a PRISMA flow diagram.

### Search strategy

Key words were generated by the authors by selecting terms commonly used in key publications from the literature (see table 1). Truncation (eg, ‘*’ or ‘$’) will be used to allow suffixes of key words to be captured during the searches (eg, ‘audiolog*’ to include ‘audiology’ and ‘audiological’). For each database, the search, using Boolean logic (eg, ‘hearing impair*’ OR ‘hearing loss’), will be split into two domains: audiological terms (search 1) and cardiovascular terms (search 2). Wherever possible, MeSH terms for each domain will be used when searching databases (see table 1). MeSH terms may vary across database searches to comply with the MeSH terminology of each database.

**Table 1.** Key words searched in titles and abstracts and Medical Subject Headings (MeSH) terms

| **Domain** | **Keywords** | **MeSH terms (for PubMed)** |
| --- | --- | --- |
| Audiological terms (search 1) | audiolog* OR audiological function* OR audiological health OR audiometry OR audiometric OR auditory OR central auditory OR cochlea* OR hearing OR hearing function* OR hearing impair* OR hearing loss OR peripheral auditory OR speech in noise OR speech identification OR speech perception OR speech intelligibility OR speech recognition | audiometry OR audiometry, pure tone OR auditory threshold OR cochlear diseases OR hearing OR hearing disorders OR hearing loss OR hearing loss, bilateral OR hearing loss, central OR hearing loss, functional OR hearing loss, high frequency OR hearing loss, noise induced OR hearing loss, sensorineural OR hearing loss, sudden OR hearing loss, unilateral OR hearing tests OR otoacoustic emissions, spontaneous OR speech OR speech perception OR speech intelligibility OR auditory pathways OR auditory pathway disorders, central |
| Cardiovascular terms (search 2) | arterial OR artery OR atherosclerosis OR blood pressure OR body mass index OR cardiovascular OR exercis* OR fitness OR physical activity OR vascular OR BMI OR arterial stiffness OR endothelial function OR microvascular OR vascular resistance OR cardiorespiratory OR pulse wave* | cardiovascular abnormalities OR cardiovascular diseases OR cardiovascular physiological phenomena OR cardiovascular system OR physical fitness OR exercise OR body mass index OR biomarkers |

After completing searches 1 and 2, Boolean logic will be used to combine the two searches (eg, ‘search 1’ AND ‘search 2’) to retrieve studies that include both domains. The strategy used for each database will be reported to ensure replicability.

### Data management

Studies retrieved from all databases will be imported into Covidence (https://www.covidence.org/) that automatically discards any duplicates (search results having the same title, year of publication, volume and author list). A manual check will be conducted to remove any studies appearing repeatedly with slight differences in their reference. Reasons for exclusion will be noted.

### Data selection process

The data selection process will be carried out by two reviewers (authors RD and CF) using Covidence. It will consist of two sifting phases. In phase 1, the title and abstract of all retrieved studies from the searches will be screened against the eligibility criteria. Studies that fit the inclusion criteria will be included in the second phase. In case that there is not enough information in the title or abstract of a study to decide if it fits the inclusion criteria, the study will also be included in phase 2. In this phase, the full text of studies will be reviewed against the eligibility criteria, and this will result in the rejection or acceptance of the study. The number of rejected and accepted studies will be presented in a PRISMA flow diagram.

### Data collection process

Studies accepted during the data selection process will be used for data collection. A data extraction form, created using Covidence, will be used to store the data extracted from each study. Each reviewer will receive clear instructions on how to conduct the data collection process to ensure all information will be collected and recorded consistently. Any discrepancies between reviewers will be decided by authors EO or LB. A minimum of 10% of the accepted studies will be used to pilot the data collection form and any changes needed to the form will be implemented prior to conducting the data collection proper on the entirety of the accepted studies.

### Data items

Items extracted from the studies will be recorded in the following way:

1. *Study information*: study identification number, author(s), year of publication, title, language of publication, publication type, study objective(s), and study design
2. *Population*: number of participants, recruitment type, ages, hearing status, CV status, geographical location, sex, and race/ethnicity
3. *Factors*: CV health measures: direct measures of CV health (eg, VO_2_ max, pulse wave velocity); indirect measures of CV health (eg, CV risk factors such as BMI, hypertension, self-reported smoking status)
4. *Outcomes*: primary - peripheral auditory functioning (eg, as measured by audiometric thresholds, OAEs) and central auditory functioning (eg, as measured by speech-identification tasks, self-reports of listening abilities); secondary - cognitive functioning
5. *Outcome results*: reported associations between CV health and auditory functions
6. *Risk of bias*: level of bias risk for each study

In case of missing data, corresponding authors of the studies in question will be contacted. Records will be kept of any data that cannot be obtained.

### Risk of bias in individual studies

The risk of bias for individual studies will be assessed by authors RD and CF using the Appraisal tool for Cross-Sectional Studies (AXIS).[72] This a 20-item questionnaire exploring separately the different sections (ie, introduction, methodology, results and discussion) of each study. Each item is rated ‘Yes’, ‘No’ or ‘Do not know/Comment’.

### Data synthesis

Using cut-offs suggested by the Cochrane library,[73] heterogeneity of the data will be explored using the *I*^2^ statistic. Homogenous data (with *I*^2^ < 50%) will be pooled for a meta-analysis, while heterogenous data will be analysed using a narrative synthesis. In the case that the entire dataset shows high levels of heterogeneity (*I*^2^ > 60%), a narrative synthesis will be conducted, following the Synthesis Without Meta-analysis (SWiM) framework.[74]

### Meta-bias(es)

If a meta-analysis is conducted, forest plots will demonstrate any meta-bias(es) that may occur.

### Confidence in cumulative evidence

The overall risk of bias will be reviewed using phases 2 and 3 of the ROBIS tool.[75] Phase 2 of the ROBIS tool has 21 questions across four domains (ie, study eligibility criteria, identification and selection of studies, data collection and study appraisal, synthesis and findings). Phase 3 is the overall judgment of bias. In addition, the overall quality of this systematic review will be reviewed using the Critical Appraisal Skills Programme Checklist for Systematic Reviews.[76] This is comprised of 10 questions, answered with ‘Yes’, ‘No’ or ‘Can’t tell’.

### Patient and public involvement

Patient and public involvement is not required for this systematic review.

## Data Availability

All data produced in the present work are contained in the manuscript

## ETHICS AND DISSEMINATION

Ethical approval is not required as no primary data will be collected. Findings of the systematic review will be reported at scientific conferences and in peer-reviewed journals following the PRISMA guidelines.

## AUTHOR CONTRIBUTIONS

CF initiated the idea for this systematic review. RD prepared the initial version of the protocol. All authors selected the eligibility criteria, key words, and the data items list. RD and CF wrote the manuscript. EO and LB provided feedback on earlier versions of the manuscript. All authors approved the final version of the manuscript.

## ACKNOWLEDGEMENTS

The authors are indebted to Nathan Rush (librarian at Loughborough University) and Natalie Pearson for helpful discussions about protocols for systematic reviews.

## DATA SHARING STATEMENT

All data collected according to the data items will be available on request to the extent that they are not included in the final published systematic review article.

